# Association of COVID-19 spread with on-demand testing

**DOI:** 10.1101/2020.12.04.20244079

**Authors:** Leon S. Robertson

**Affiliations:** Yale University, 60 College St., New Haven, CT

## Abstract

Comparisons of COVID-19 testing policies in 99 countries indicate that testing on demand is associated with an increase in cases 14 days after the tests. Adjusted for number of new cases and the positivity rate, for each 10,000 negative tests on a given day there were 90-110 new cases in 14 days that would not have otherwise occurred. Approximately 3.1 million or 21 percent of new cases in periods of on-demand testing are likely due to that policy through early November 2020. During periods in a given country when only persons with symptoms were tested, or persons with symptoms and key vulnerable populations were tested, negative tests were associated with fewer new cases 14 days out. Apparently when tests are available on demand, those who test negative are engaging in activities that increase the risk of exposure.

## Introduction

Public policy responses to the COVID-19 pandemic varied among countries and, in some cases, among provinces, states and cities within countries. As biological tests of infection of individuals by the virus became available, countries that used tests to identify cases adopted one of three testing policies: 1. Test persons who present with symptoms, 2. Test persons with symptoms plus populations vulnerable to more severe outcomes such as people in nursing homes, the elderly generally, and persons with preexistent conditions known to exacerbate severity of the disease. 3. Test on demand including free testing in some countries. The justification for the latter policy was to identify asymptomatic individuals who could nevertheless spread the virus to others as well as adjust other policies such as school and business closings based on the proportion positive of those tested. Tracing the contacts of those who tested positive and urging or requiring all who tested positive to quarantine themselves for 14 days would theoretically reduce the spread of the virus. Some countries changed testing policies as well as other policies from time to time.

On-demand testing does not necessarily indicate the true positivity rate in a population because the people who self-select to be tested are not necessarily representative of the population. Furthermore, if enough people who receive a negative test behave in such a way as to increase their risk of exposure to the virus, the test-on-demand policy could result in increased spread of the virus. This hypothesis came to mind as news reports indicated long lines of people waiting to be tested prior to the Thanksgiving holiday in the U.S., a holiday traditionally marked by huge increases in travel to large family gatherings. Although public health officials issued repeated warnings that a negative test did not mean that one could not get the virus after the test and spread it to others, it appeared that a large number of people were ignoring the warnings and planning to travel if they received a negative test.

The purpose of this paper is to report differences in spread of COVID-19 during times when countries were testing only the symptomatic, the symptomatic plus the more vulnerable, or allowing testing on demand.

## Materials and Methods

Seven-day moving averages of daily numbers of tests, positivity rates and numbers of new cases among 99 countries where the data were available as of November 21, 2020 were downloaded from ourworldindata.org (https://ourworldindata.org/coronavirus-testing). Hasell et al. ^1^ provide a description of the database and derived variables as of August 31, 2020. The testing policy on a given day in each country was also downloaded from ourworldindata.org and was matched by country and date to the file containing tests and cases.

Since symptoms in the infected may occur as much as two weeks after exposure, a least squares regression model was used to predict cases two weeks after a given day that a specified number of tests, cases and positivity occurred. The form of the model is:

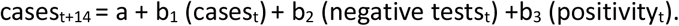

The number of negative tests on a given day was obtained by multiplying the positivity rate times the number of new tests on a given day and subtracting the result from the total number of tests. Controlling for cases at time t adjusts for the stage of the pandemic at a given time in a given country. If negative tests result in increased behavior that exposes the individual to risk of infection, then b2 will be positive, indicative of an increase in cases two weeks out in relation to negative tests. The regression coefficients were estimated separately for each of the testing policies.

## Results

The regression coefficients and 95 percent confidence intervals are presented in Table 1.

**Table 1.**
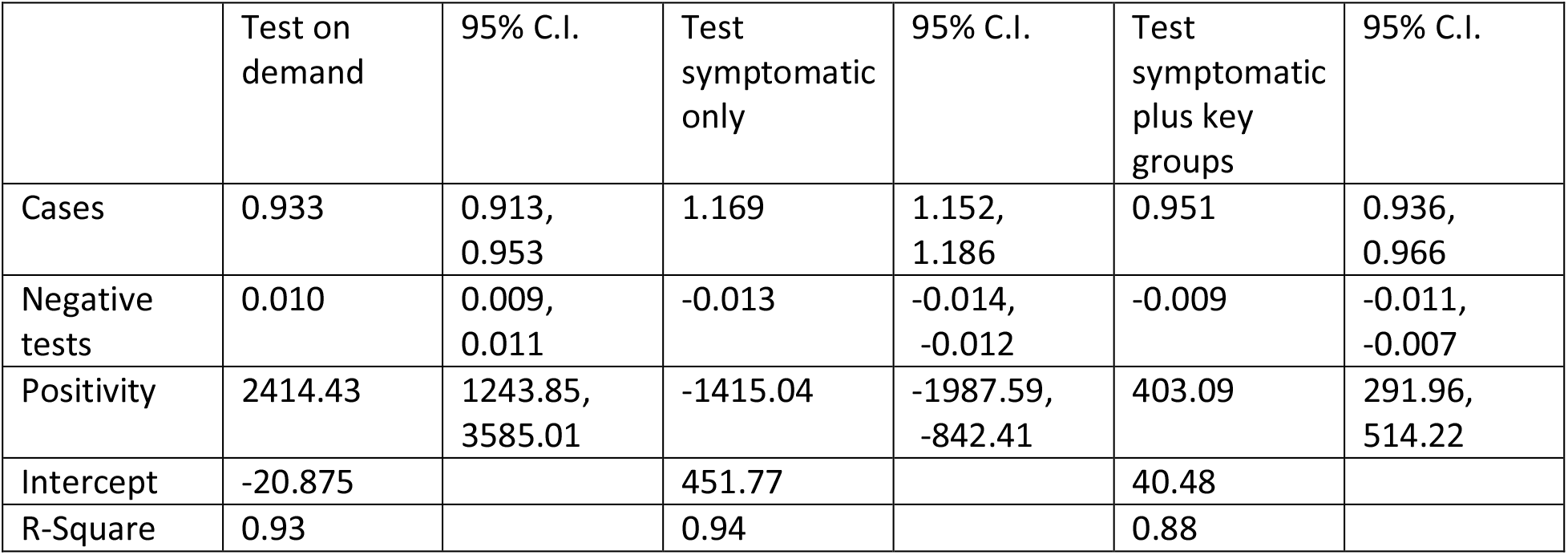
New Cases in 14 Days Predicted by Number of New Cases Extant, Negative Tests, and Positivity on a Given Day

The 0.01 positive coefficient on negative tests during times of unlimited testing indicates that for each 10,000 tests on a given day there will be 90-110 new cases in 14 days that would not have otherwise occurred. The negative coefficients for days that only the symptomatic or the symptomatic and key groups were tested indicate the opposite, about a hundred fewer cases for each 10,000 tested in 14 days than would have occurred otherwise. There were 308.5 million negative tests in on-demand testing periods in the studied countries and 14.6 million cases. At 100 new cases per 10,000 tests, approximately 3.1 million or 21 percent of new cases in periods of on-demand testing are likely due to post-test behavior.

An increment of .01 in positivity predicts 12-35 new cases in 14 days when testing is on demand and about 4 new cases in 14 days when the symptomatic and key groups are tested. About 14 fewer cases in 14 days are predicted by a .01 increment in positivity when only the symptomatic are tested.

## Discussion

This important dataset is missing a key piece of information: the number of tests in each policy group that were the result of tracing contacts of persons who tested positive. Presumably countries with symptoms only and symptoms plus key group testing did tracing and found contacts who tested negative in a proportion of cases. In the U.S. with an on-demand testing policy, tracing varied among the states and in many instances the number of cases overwhelmed tracer personnel to the point that tracing effectiveness was compromised. Some people, when traced, refused to be tested and some of those who tested positive refused to self-quarantine.^3^ The limited supply of testing materials reduced the numbers tested in some areas. Nursing organizations complained that health care employers were not testing their members while professional and college sports teams were tested frequently. ^4^

Expensive and unrepresentative mass testing is not necessary to detect the spread of COVID-19 in a community. The trajectory of the curve of new cases is likely to continue if measures to limit interpersonal contact and increase wearing of masks are not adopted. ^5^ Also, tests of virus concentrations in community waste water have been shown to predict the spread. ^6,7^ Research on U.S. state and local rules regarding mandatory mask use and restrictions on gatherings indicates that these policies reduce COVID-19 mortality rates. ^8^

## Data Availability

The data are available at: https://ourworldindata.org/coronavirus-testing

